# Influence of Factor V Leiden mutation and Protein C/S deficiencies on preeclampsia among Sudanese women

**DOI:** 10.1101/2024.10.31.24316543

**Authors:** Faris Abdon, Maha Elamin, Khalid Hussein Bakheit

## Abstract

**Background:** Preeclampsia (PE) is a serious pregnancy complication that poses significant health risks to both mothers and babies. Genetic factors like thrombophilia mutations and deficiencies in natural anticoagulants might contribute to its development, but their exact roles are not well understood, especially among Sudanese women.

**Objective:** To assess the relationship between the Factor V Leiden (FVL) thrombophilic mutation and reduced levels of natural anticoagulants Protein C (PC) and Protein S (PS) with the occurrence of PE among Sudanese women.

**Methods and materials:** We conducted a case-control study that included a total of 300 women, divided equally into three groups: 100 with PE, 100 healthy pregnant women, and 100 healthy non-pregnant women. To detect FVL mutations, we used PCR-RFLP analysis. Levels of PC and PS were measured using colorimetric assays. We applied logistic regression analyses to assess the relationships between these variables and the risk of developing PE.

**Results:** Our findings showed no significant link between FVL mutations and PE (p=.390). PC levels on their own did not emerge as significant independent predictors of PE (OR 1.01, 95% CI 0.99–1.02, p=.419). However, women with low PC and S levels were strongly associated with PE in both univariate and multivariate analyses (OR 77.67, 95% CI 8.97– 672.5, p<.001). This combination was significantly more common in the PE group than in the control group (p<.001). Additionally, reduced PS levels were significantly associated with an increased risk of PE.

**Conclusion:** Combined PC/PS deficiencies are strongly associated with PE among Sudanese women, indicating a significant role of these natural anticoagulants in the disease’s pathogenesis. FVL mutation was not significantly linked with PE in this population.

## Introduction

Thrombophilia, or an increased tendency to form blood clots, can be due to inherited or acquired factors. Pregnancy naturally elevates the likelihood of clotting, and when combined with thrombophilia, the chance of thrombosis becomes higher (1). Mutations that result in protein deficiencies, such as PC or PS (loss-of-function mutations), or mutations that increase the activity of clotting factors, such as FVL (gain-of-function mutations), can cause inherited thrombophilia. Despite some research indicating a correlation between inherited thrombophilia and preeclampsia (PE), this association remains a subject of debate (1).

The most prevalent inherited thrombophilia is FVL, which is caused by a specific mutation (G1691A) in the Factor V gene. This mutation causes an amino acid change (Arg506Gln) in Factor V (f5), rendering it resistant to inactivation by activated PC. This resistance results in a hypercoagulable state, particularly during pregnancy (2-4). The FVL mutation has been linked to an elevated risk of PE and pregnancy loss (5, 6). Meta-analyses have verified a substantial correlation between the FVL mutation and hypertensive disorders of pregnancy, indicating that FVL may be a genetic risk factor for PE (6). Nevertheless, the correlation between PE and FVL remains a topic of debate. Some studies indicate that PE patients have a higher prevalence of thrombophilic disorders than controls; however, they do not establish a direct correlation between FVL and PE (7, 8). For example, a study on the Kolar population found no significant association between the FVL mutation and PE (9), and others suggest that due to inconsistent findings, routine thrombophilia screening during pregnancy may not be justified (8, 10). These findings highlight the importance of considering population-specific genetic backgrounds when studying the genetic factors contributing to PE (4, 10).

Genetic predispositions that may contribute to PE among Sudanese women have been identified in research conducted in Sudan. For example, a study discovered a substantial correlation between PE and the FVL G1691A mutation. FVL was detected in 9.6% of preeclamptic women, while it was present in only 0.6% of controls (11). Another study confirmed a significant association between FVL mutations and severe PE, with a mutation prevalence of 16% in cases compared to 0% in controls (12).

PS and PC are essential components of the body’s anticoagulant system, and their deficiencies are recognized to elevate the likelihood of thrombotic events (13, 14). PC is a vitamin K-dependent enzyme that is crucial for the regulation of blood coagulation. It is activated by the thrombin-thrombomodulin complex and, in conjunction with PS, prevents the formation of thrombin by deactivating coagulation cofactors (13). PS, also a vitamin K-dependent glycoprotein, serves as a cofactor for activated PC and has independent anticoagulant properties (13). PS and PC levels decrease during pregnancy, and any deficiencies can result in a hypercoagulable state, which may contribute to the development of PE (4).

The prevalence and impact of PC and S deficiencies in PE have been the subject of conflicting results in research. For instance, Igwe et al. reported that Nigerian women with increased pregnancy loss had reduced levels of PC, whereas other studies did not observe any significant differences in PS levels between preeclamptic and healthy women (15, 16).

PE and other adverse outcomes are significantly increased by consanguinity, notably marriages between cousins. In Lahore, Pakistan, consanguinity was associated with 50% of PE and 70% of eclampsia cases (17). This practice, common in the Middle East due to sociocultural reasons, also increases the risk of congenital disorders (18). However, in Iran, consanguineous marriage was linked to an increased risk of stillbirth, particularly preterm stillbirths (19). Additionally, PE tends to be more severe in consanguineous couples, with higher blood pressure and proteinuria, suggesting a genetic predisposition (20).

## Methods and materials

From 2019 to 2021, this case-control study was conducted at Omdurman Maternity Hospital in Khartoum State, Sudan. The objective was to study the correlation between PE and FVL mutations in Sudanese women and to investigate natural blood coagulation inhibitors Activated Protein C Resistance (APC-R). The study comprised 300 women, who were equally distributed among three groups: 100 women diagnosed with PE, 100 healthy expectant women, and 100 healthy non-pregnant women.

The PE group consisted of women who had blood pressure readings that exceeded 140/90 mmHg on two or more occasions after 20 weeks of gestation and demonstrated significant proteinuria (>300 mg/dl or 2+ on dipstick examinations). The control group of healthy expectant women consisted of normotensive women with uncomplicated singleton pregnancies, and their delivery dates were closely matched to those of the preeclamptic group. The control group of healthy non-pregnant women consisted of women who were non-smokers, did not have a history of thromboembolic diseases, and were not taking anticoagulant medications.. Inclusion criteria for the study required that participants meet the specific conditions outlined for each group. Exclusion criteria applied to all groups included a history of chronic hypertension, diabetes, kidney disease, autoimmune disorders, smoking, thromboembolic events, and anticoagulant drugs. Additionally, PE group excluded women with preexisting hypertension before 20 weeks of gestation, multiple pregnancies (such as twins or more), and other systemic health conditions that could confound the study results.

### Data Collection and Biochemical Assays

Demographic, clinical, and biochemical information was gathered from each participant using a validated questionnaire. Venous blood samples were collected to extract genomic DNA and assess PC and S levels. PC was activated with a chromogenic substrate, and the absorbance was measured spectrophotometrically to ascertain PC levels. Similarly, PS concentrations were determined by initiating a reaction with thrombin and a chromogenic substrate, which resulted in a change in color intensity that indicated the concentration of PS. PC and PS were assessed for their activity levels, with reference ranges of 65%–150% for PC and 57%–131% for PS (ABIM. Laboratory Tests Reference Ranges. Revised January 2024. Available from: https://www.abim.org) (21).

### Molecular Methods

The G-DEXTM IIb Genomic DNA Extraction Kits were employed to isolate DNA from venous blood samples, and the extracted DNA was quantified using a NanoDrop spectrophotometer. PCR-RFLP was employed to conduct Restriction Fragment Length Polymorphism (RFLP) analysis in order to identify missense mutations in the F5 gene, specifically the FVL (R506Q) polymorphism. Specific primers were employed to target a 206 base pair (bp) region of the FVL gene during PCR amplification. At 95°C for 10 minutes, the PCR protocol initiated with an initial activation. Subsequently, 35 cycles of denaturation at 95°C for 30 seconds, annealing at 55°C for 1 minute, and extension at 70°C for 3 minutes were conducted. Following amplification, the PCR products were digested with the MnII restriction enzyme and separated by electrophoresis on a 1.5% agarose gel. The DNA fragments were subsequently observed by visualizing the gel under UV light. Genotypes were determined based on the pattern of fragments: two fragments (47 and 159 bp) indicated a mutant homozygous genotype (A/A), three fragments (136, 47, and 23 bp) indicated a wild-type homozygous genotype (G/G), and four fragments (34, 47, 23, and 102 bp) showed a heterozygous genotype (A/G).

### Ethics methods

This study was approved by the Ethics Committee of Al-Neelain University (IRB Serial No: NU-IRB-18-8-8-41) on (30/08/2018). All participants provided written informed consent prior to their inclusion in the study. Informed consent was obtained using standardized consent forms, signed and dated by each participant to ensure proper documentation and confidentiality. The research was conducted in accordance with the Declaration of Helsinki and adhered to all relevant institutional and national ethical guidelines.

### Data analysis

In order to identify PE predictors among Sudanese women, data analysis was performed using IBM SPSS Statistics version 29.0. The sociodemographic, clinical, and biochemical characteristics of the case and control groups were compared using descriptive statistics. Categorical variables were analyzed using chi-square tests, while continuous variables were analyzed using t-tests or Mann-Whitney U tests, as appropriate. In order to identify prospective predictors of PE, univariate logistic regression was implemented. Subsequently, multivariate logistic regression was implemented to mitigate any potential confounding variables. A backward incremental method was employed to identify significant predictors. The logistic regression model’s ability to discriminate between cases and controls was evaluated using the Hosmer-Lemeshow test, and the area under the Receiver Operating Characteristic (ROC) curve (AUC) was calculated to assess its goodness-of-fit. Furthermore, the analysis was not significantly affected by any significant issues, as evidenced by the assessment of multicollinearity using variance inflation factors (VIFs).

## Results

The median age of women with PE was considerably higher (36 years) than that of healthy pregnant women (29 years) and non-pregnant women (31 years) (Table 1), as evidenced by a p-value of less than .001. The non-pregnant group (72%) had a higher prevalence of secondary education than the PE group (58%) and the healthy expectant group (51%), with p-values of .038 and .002, respectively.

**Table 1.** Comparison of Sociodemographic, Clinical, and Laboratory Parameters Among PE Cases, Healthy Pregnant Controls, and Non-Pregnant Controls.

|  | Cases<br>(N=100) | Controls<br>(N=200) |  |  |
| --- | --- | --- | --- | --- |
| Characteristics | (1)<br>Preeclampsia<br>Group | (2)<br>Healthy<br>Pregnant Group | (3)<br>Non-Pregnant<br>Group | p-value |
| Age in years<br>median (IQR) | 36 (31.25-39) | 29 (27-32) | 31 (27-34) | 1vs.2: <.001 |
|  |  |  |  | 2vs.3: .368 |
|  |  |  |  | 1vs.3: <.001 |
| Educational Level<br>(Up to Secondary %) | 58% | 51% | 72% | 1vs.2: .320 |
|  |  |  |  | 2vs.3: .002 |
|  |  |  |  | 1vs.3: .038 |
| Employment<br>(Employed %) | 23% | 30% | 38% | 1vs.2: .262 |
|  |  |  |  | 2vs.3: .232 |
|  |  |  |  | 1vs.3: .021 |
| Consanguinity | 67% | 65% | 20% | 1vs.2: .765 |
|  |  |  |  | 2vs.3: <.001 |
|  |  |  |  | 1vs.3: <.001 |
| Contraceptive Pills | 13% | 9% | 1% | 1vs.2: .366 |
|  |  |  |  | 2vs.3: .009 |
|  |  |  |  | 1vs.3: <.001 |
| Presenting Blood Pressure |  |  |  |  |
| Systolic BP Median-IQR | 150 (140 - 160) | 120 (113 - 120) | 120 (115-120) | 1vs.2: <.001 |
|  |  |  |  | 2vs.3: .392 |
|  |  |  |  | 1vs.3: <.001 |
| Diastolic BP Median-IQR | 100 (90 - 120) | 80 (70 - 80) | 80 (80-80) | 1vs.2: <.001 |
|  |  |  |  | 2vs.3: .080 |
|  |  |  |  | 1vs.3: <.001 |
| Platelets Median-IQR | 244 (233.3-252.8) | 277.5 (184 - 312) | 290 (197.5-357.5) | 1vs.2: .006 |
|  |  |  |  | 2vs.3: .409 |
|  |  |  |  | 1vs.3: .011 |
| Levels of PC and PS |  |  |  |  |
| PC Median-IQR | 107.8 (101.8-117.2) | 102.0 (97 - 109) | 88.5 (78.3-105.0) | 1vs.2: <.001 |
|  |  |  |  | 2vs.3: <.001 |
|  |  |  |  | 1vs.3: <.001 |
| PS Median-IQR | 77.4 (50.2 - 88.5) | 49.3 (43.2 – 60.8) | 77.5 (66.2-96.6) | 1vs.2: <.001 |
|  |  |  |  | 2vs.3: <.001 |
|  |  |  |  | 1vs.3: .004 |

The employment rates of the PE group were 23% lower than those of the non-pregnant group (38%; p=.021). However, there was no significant difference between the PE group and the healthy expectant group. Contrary to the non-pregnant group (20%), consanguinity was more prevalent in the PE group (67%) and the healthy pregnant group (65%) (p<.001). However, there was no significant difference between the PE and healthy pregnancy groups.

The use of contraceptive medications was significantly higher in the PE group (13%) than in the non-pregnant group (1%) (p<.001), and it differed between the healthy pregnant group (9%) and the non-pregnant controls (p=.009). The PE group exhibited a higher prevalence of previous pregnancy complications than both the healthy expectant and non-pregnant groups (p<.001 and p=.010).

The PE group exhibited substantially higher systolic and diastolic blood pressures than either of the control groups (p<.001). The platelet counts of the PE group were significantly lower than those of the healthy pregnant and non-pregnant groups (p=.006 and p=.011, respectively).

The PE group exhibited the maximum PC levels, followed by the healthy pregnant group and the non-pregnant group (p<.001). Conversely, the non-pregnant group exhibited the highest PS levels, with the PE group exhibiting levels that were higher than those of the healthy expectant group (p<.001).

FVL (R506Q) genotypes distribution was analyzed using PCR-RFLP and visualized with agarose gel electrophoresis, as shown in (Fig 1). Homozygous mutant (AA) individuals had two DNA fragments (47 and 159 bp), wild-type homozygous (GG) individuals had three fragments (136, 47, and 23 bp), and heterozygous (AG) individuals had four fragments (34, 47, 23, and 102 bp).

**Fig 1.**
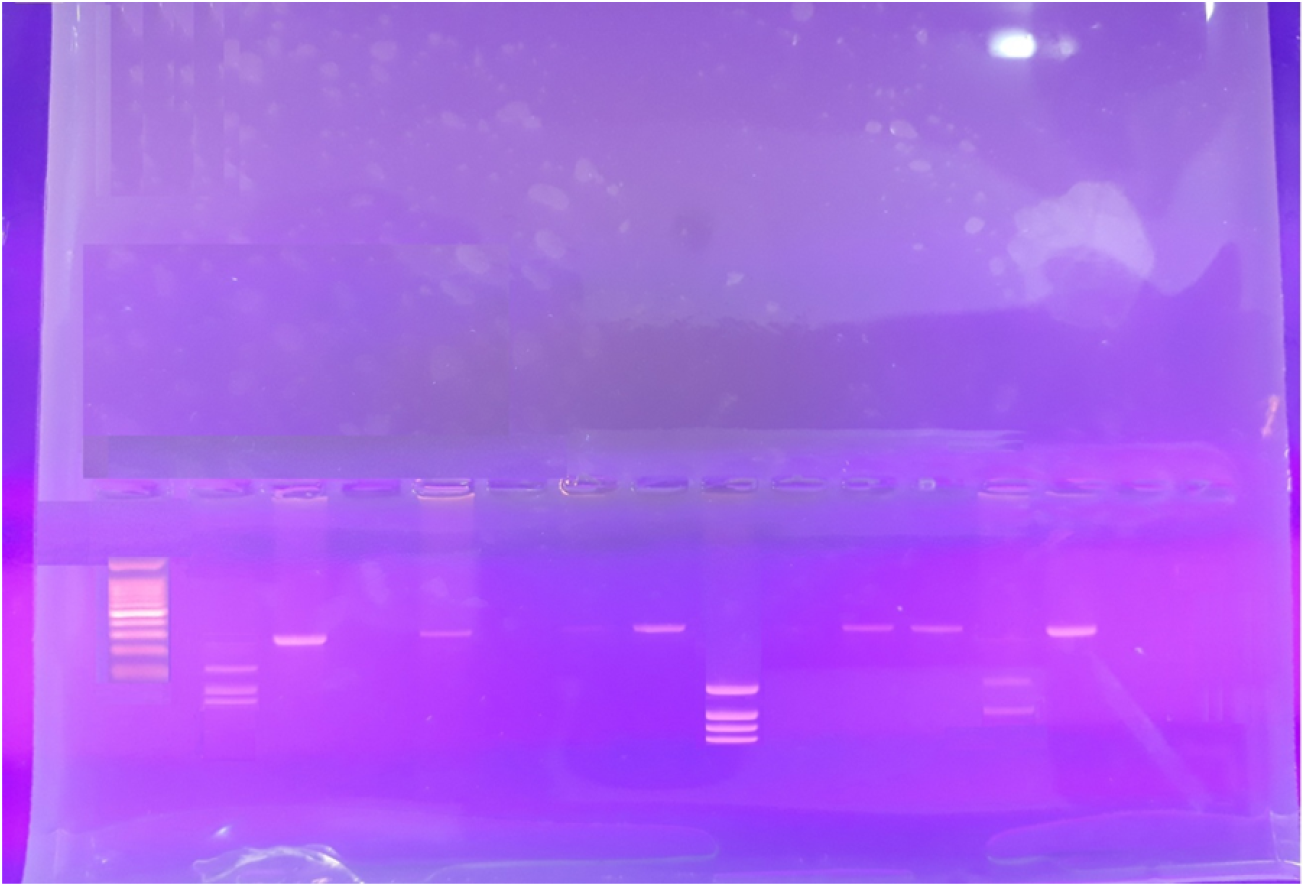
PCR-RFLP Analysis of FVL (R506Q) Genotypes by Agarose Gel Electrophoresis

Although these differences were not statistically significant, the PE group had a higher percentage of homozygous positive (AA) individuals (8%) than the healthy expectant and non-pregnant groups (3% each), as indicated by (Table 2).

**Table 2.** Distribution of FVL Genotypes and Alleles Among Study Groups.

| Genotype | Preeclampsia<br>(N=100) | Healthy<br>Pregnant<br>(N=100) | Non-Pregnant<br>(N=100) | Total<br>(N=300) |
| --- | --- | --- | --- | --- |
| Homozygous Positive FVL / AA | 8% | 3% | 3% | 14 (4.67%) |
| Heterozygous FVL / GA | 0 | 2% | 2% | 4 (1.33%) |
| Negative FVL / GG | 92% | 95% | 95% | 282 (94%) |
| Allele A | 16 | 8 | 8 | 32 |
| Allele G | 184 | 192 | 192 | 568 |

The distribution of thrombophilia markers among the study groups is illustrated in Table 3. The FVL mutation was prevalent in all groups, with no evidence of significant differences. The PE group exhibited a substantially higher prevalence of low PC compared to the healthy pregnant group (p<.001), while the non-pregnant group exhibited a moderate difference (p=.009). The PE group exhibited a significantly higher frequency of low PS compared to the healthy pregnant group (p=.003), and the healthy pregnant group exhibited a significantly higher frequency of low PS compared to the non-pregnant group (p<.001). Furthermore, the proportion of reduced PS in the PE group was significantly higher than that in the non-pregnant group (p=.039).

**Table 3:** Distribution of Thrombophilia Markers Among PE Cases, Healthy Pregnant Controls, and Non-Pregnant Controls.

|  | Cases<br>(N=100) | Controls (N=200) |  |  |
| --- | --- | --- | --- | --- |
|  | (1)<br>Preeclampsia<br>Group | (2)<br>Healthy<br>Pregnant Group | (3)<br>Non-Pregnant<br>Group | p-value |
| Positive FVL | 8 | 5 | 5 | 1vs.2: .390<br>2vs.3: >.999<br>1vs.3: .390 |
| Low PC | 11 | 1 | 9 | 1vs.2: <.001 |
|  |  |  |  | 2vs.3: .009 |
|  |  |  |  | 1vs.3: .637 |
| Low PS | 31 | 65 | 1 | 1vs.2: .003 |
|  |  |  |  | 2 vs.3 <.001 |
|  |  |  |  | 1vs.3: .039 |
| Combination of (Positive FVL + Low PS) | 1 | 1 | 0 | 1vs.2: >.999 |
|  |  |  |  | 2vs.3: .316 |
|  |  |  |  | 1vs.3: .316 |
| Combination of (Low PC + Low PS) | 10 | 1 | 0 | 1vs.2: .005 |
|  |  |  |  | 2vs.3: .316 |
|  |  |  |  | 1vs.3: .001 |
| Combination of (Positive FVL + Low PC + Low PS) | 1 | 0 | 0 | 1vs.2: .316 |
|  |  |  |  | 2vs.3: N/A |
|  |  |  |  | 1vs.3: .316 |
| Combination of (Negative FVL, Normal PC, and Normal PS) | 62 | 31 | 85 | 1vs.2: <.001 |
|  |  |  |  | 2vs.3: <.001 |
|  |  |  |  | 1vs.3: <.001 |

Significantly more frequently in the PE group than in the healthy pregnant group (p=.005) and the non-pregnant group (p=.001) was the combination of low PC and low PS. The PE group was the sole location of the rare combination of low PC, low PS, and positive FVL; however, the number of cases was insufficient to detect significant differences. In contrast, the PE group exhibited a substantially lower prevalence of the combination of negative FVL with normal PC and PS compared to both control groups (p<.001). This suggests that women with PE have a unique thrombophilia profile.

The relationship between thrombophilia markers, clinical variables, and the occurrence of PE was investigated using logistic regression analyses, as illustrated in Table 4. PE was not significantly associated with consanguinity (p=.765) or the use of oral contraceptive tablets (p=.368) in the univariate analysis. PS levels were significantly correlated with PE in both univariate (OR 1.07, 95% CI 1.04–1.09, p<.001) and multivariate analyses (OR 1.09, 95% CI 1.06–1.11, p<.001).

**Table 4.** Thrombophilia Markers and Clinical Variables in PE: Univariate and Multivariate Analysis vs. Healthy Pregnant Controls

| Variable | Univariate OR(95% CI) | Univariate p value | Multivariate aOR (95% CI) | Multivariate p-value |
| --- | --- | --- | --- | --- |
| Consanguinity | 1.09(0.61-1.96) | .765 | — | — |
| Oral Contraceptive Pills | 1.51(0.62-3.71) | .368 | — | — |
| **PC and PS** |  |  |  |  |
| PC Levels | 1.01(0.99-1.02) | .419 | — | — |
| PS Levels | 1.07(1.04-1.09) | <.001 | 1.09(1.06-1.11) | <.001 |
| Combination of Low PC + Low PS | 11.00(1.38-87.61) | .024 | 77.67(8.97-672.5) | <.001 |
| **FVL** |  |  |  |  |
| Positive FVL (Homo and Heterozygous) | 1.65(0.52-5.24) | .394 | — | — |
| Negative FVL | Reference |  |  |  |

The combination of low PC and low PS was significantly associated with PE, as evidenced by the increased risks observed in both univariate (OR 11.00, 95% CI 1.38–87.61, p=.024) and multivariate analyses (OR 77.67, 95% CI 8.97–672.5, p<.001) analyses. This suggest a synergistic effect. PE did not exhibit a significant association with the positive FVL status (p=.394).

## Discussion

In this study, the role of natural blood clotting inhibitors, specifically PC and PS, as well as the genetic polymorphism FVL, in preeclampsia (PE) among Sudanese women was examined.

Within this study, homozygous FVL was more common in the PE group (8%) than in the control groups (5%), although the difference was not statistically significant (p=.390). Our results are in agreement with a prior Sudanese study by Ahmed et al., which found a 9.6% prevalence of FVL among women with PE (11). Globally, there was a great inconsistency in FVL rates, 0.76%, 8%, 15.4%, and 33% in Nigeria, Turkey, Egypt, and Ghana respectively. (22),(23),(24) The prevalence of FVL in our control group was (5%) which is consistent with findings in regions such as Saudi Arabia (4.4%) (25), Egypt (2.5-10.2%), Tunisia (3.0-13.6%), Turkey (4.6-9.8%) (26), and Europe (2-15%) (27), the USA (3.2-6%), and Caucasians (3-8%). (26), (28) Although FVL was more prevalent in the PE group, it was not significantly associated with the condition in both univariate and multivariate analyses. This lack of significant association suggests that FVL may not be a major factor in PE within this multiethnic population, potentially due to the intricate interactions between genetic and environmental factors that diminish its impact (4, 29). Similarly, Changavala, Deveer, Madkhaly, and Vicoveanu reported similar findings but did not find a significant link between FVL and PE (9, 30-32). However, other studies by Ahmed, Akhtar, and Alsheikh have indicated a potential association between FVL and PE in specific populations (11, 23, 33), Emphasizing the population-dependent nature of this relationship.

PC levels were substantially higher in the PE group (11%) than in healthy pregnant controls (1%, p<.001). However, there was no significant difference between the PE group and non-pregnant controls (9%, p=.637). The median PC levels were also lower in the PE group than in both control groups (p<.001). PC was not identified as an independent predictor of PE by logistic regression (OR 1.01, 95% CI 0.99-1.02, p=.419). This is in accordance with the results of Katz et al., who demonstrated that PC activity remains constant during pregnancy. (34). Research by Saleh et al., Okoye et al., and Mitriuc et al. also reported reduced PC in PE but suggested that PC deficiency alone is not a strong predictor due to the complex interplay of coagulation factors during pregnancy (35-37). This highlights the importance of considering multiple clotting factors when assessing the risk of PE.

Previous studies have suggested that low PS levels during pregnancy represent a normal physiological adaptation, particularly in the later stages of gestation (34-36, 38). Our findings showed that healthy pregnant women had lower PS levels than preeclamptic women. The reasons for this discrepancy are unclear but may involve complex physiological or genetic factors unique to our study population. Additionally, the genetic diversity of the Sudanese population may contribute to these variations in PS levels and their association with PE.

The PE group exhibited a substantially higher prevalence of the combination of low PC and low PS compared to both control groups (p<.001). Consequently, this combination was a strong predictor of PE in both univariate and multivariate analyses. While pregnancy typically enhances blood clotting to prevent hemorrhage during delivery, this adaptation can increase the risk of blood clots in conditions such as PE, where clotting factors are disrupted. The increased risk of thrombosis observed in our study is corroborated by the stable role of PC as an anticoagulant and the decrease in PS levels. (34). Studies (35, 39) further emphasized the importance of measuring these proteins in states of increased blood clotting, while Jung et al. linked decreased PS activity with hypertensive disorders of pregnancy (40). These studies support our findings that combined deficiencies in PC/PS indicate a higher risk of PE.

On the other hand, consanguinity (marriage between relatives) and contraceptive use were not significant predictors of PE in this study. While consanguinity was prevalent in both groups, particularly among those with PE, the difference was not statistically significant. This is consistent with previous research (41), which also found no significant association between consanguinity and PE. However, studies (17, 20, 42) Khidri, Akhlaq, and Akram suggest that consanguinity may increase genetic risks for PE through the inheritance of recessive alleles. Population-specific factors may influence the role of consanguinity, potentially affecting its interaction with genetic and environmental influences in the development of PE.

Similarly, the use of hormonal contraceptives did not appear to be a significant risk factor for PE in this study, even though it was more prevalent among women with PE. This observation is consistent with previous research (43), which found no significant association between oral or injectable contraceptive use and PE. Although shared risk factors such as hormonal metabolism and the renin-angiotensin-aldosterone system have been suggested (44), the precise relationship between contraceptive use and hypertensive disorders in pregnancy remains unclear.

## Conclusion

This study discovered that combined deficiencies in PC and PS levels are strongly associated with a higher risk of PE among Sudanese women, significantly elevating the risk beyond individual deficiencies. Consanguinity and contraceptive use did not emerge as significant predictors of PE within our study population. The FVL mutation showed no significant relationship with PE in these women. These findings indicate that deficiencies in natural anticoagulants are crucial in the development of PE among Sudanese women.

## Data Availability

All relevant data are within the manuscript and its Supporting Information files.

## Acknowledgments

We express our heartfelt gratitude to all the women who participated in the study.

## Conflict of Interest

The authors declare that they have no conflict of interest regarding this article. They have no financial or personal relationships with other people or organizations that could inappropriately influence (bias) their work.

## Data Availability

The data supporting this study’s findings are available from the corresponding author, Dr. Faris Abdon, upon reasonable request. However, the data are not publicly available due to privacy or ethical restrictions. For any inquiries or requests, please contact.

## Notes

### Competing Interest Statement

The authors have declared no competing interest.

### Funding Statement

The author(s) received no specific funding for this work.

### Author Declarations

The study adhered to ethical guidelines, ensuring confidentiality and informed consent. The Ethics Committee of Al-Neelain University granted ethical approval (IRB Serial No: NU-IRB-18-8-8-41)

## References

1. Hotoleanu C. Thrombophilia in Pregnancy. International Journal of Cardiovascular Practice. 2019;4(1):1–6.

2. Ababio GK, Adu-Bonsaffoh K, Abindau E, Narh G, Tetteh D, Botchway F, et al. Effects of factor v Leiden polymorphism on the pathogenesis and outcomes of preeclampsia. BMC Med Genet. 2019;20(1):189.

3. Hiltunen LM, editor Factor V Leiden as risk factor for pregnancy complications : Epidemiological study of Finnish women 2011.

4. Padda J, Khalid K, Mohan A, Pokhriyal S, Batra N, Hitawala G, et al. Factor V Leiden G1691A and Prothrombin Gene G20210A Mutations on Pregnancy Outcome. Cureus. 2021;13(8):e17185.

5. Kujovich JL. Factor V Leiden thrombophilia. Genet Med. 2011;13(1):1–16.

6. Li Y, Ruan Y. Association of hypertensive disorders of pregnancy risk and factor V Leiden mutation: a meta-analysis. Journal of Obstetrics and Gynaecology Research. 2019;45(7):1303–10.

7. Saghafi N, Vatanchi AM, Tara F, Pourali L, Dadgar S. Evaluation of selected thrombotic factors among pregnant women with preeclampsia and normal pregnant women. Iranian Journal of Reproductive Medicine. 2014;12(12):793.

8. Roozbeh N, Banihashemi F, Mehraban M, Abdi F. Potential role of Factor V Leiden mutation in adverse pregnancy outcomes: an updated systematic review. Biomedical Research and Therapy. 2017;4(12):1832–46.

9. Changalvala K, Kotur P, Shetty M, Kumar KP, Jagadish T, Balakrishna S, et al. Maternal Factor V Leiden Mutation in Preeclampsia: A Case-Control South Eastern Indian Tertiary Care Hospital Based Study. Journal of Clinical & Diagnostic Research. 2020;14(2).

10. Salimi S, Saravani M, Yaghmaei M, Fazlali Z, Mokhtari M, Naghavi A, et al. The early-onset preeclampsia is associated with MTHFR and FVL polymorphisms. Archives of gynecology and obstetrics. 2015;291:1303–12.

11. Ahmed NA, Adam I, Elzaki SEG, Awooda HA, Hamdan HZ. Factor-V Leiden G1691A and prothrombin G20210A polymorphisms in Sudanese women with preeclampsia, a case-control study. BMC medical genetics. 2019;20:1–5.

12. Elzein HO, Saad AA, Yousif AA, Elamin E, Abdalhabib EK, Elzaki SG. Evaluation of Factor V Leiden and prothrombin G20210A mutations in Sudanese women with severe preeclampsia. Curr Res Transl Med. 2020;68(2):77–80.

13. Khider L, Gendron N, Mauge L. Inherited Thrombophilia in the Era of Direct Oral Anticoagulants. Int J Mol Sci. 2022;23(3).

14. Liatsikos SA, Tsikouras P, Manav B, Csorba R, von Tempelhoff GF, Galazios G. Inherited thrombophilia and reproductive disorders. J Turk Ger Gynecol Assoc. 2016;17(1):45–50.

15. Igwe C, Adias T, Eze E, Nwachuku E. Assessment of Protein C and Protein S of Pregnancy Loss Victims. International Blood Research & Reviews. 2019:1–7.

16. Dehkordi MA, Soleimani A, Haji-Gholami A, Vardanjani AK, Dehkordi SA. Association of Deficiency of Coagulation Factors (Prs, Prc, ATIII) and FVL Positivity with Preeclampsia and/or Eclampsia in Pregnant Women. Int J Hematol Oncol Stem Cell Res. 2014;8(4):5–11.

17. Akhlaq M, Khalid M, Nagi A. CONSANGUINITY AND PRESENCE OF PRE-ECLAMPSIA AND ECLAMPSIA IN A TERTIARY CARE HOSPITAL IN LAHORE.

18. Oniya O, Neves K, Ahmed B, Konje JC. A review of the reproductive consequences of consanguinity. European Journal of Obstetrics & Gynecology and Reproductive Biology. 2019;232:87–96.

19. Maghsoudlou S, Cnattingius S, Aarabi M, Montgomery SM, Semnani S, Stephansson O, et al. Consanguineous marriage, prepregnancy maternal characteristics and stillbirth risk: a population-based case–control study. Acta obstetricia et gynecologica Scandinavica. 2015;94(10):1095–101.

20. Akram W, Al Defer F. Preeclampsia and consanguinity. Mustansiriya Medical Journal. 2017;16(3):1–10.

21. ABIM. Laboratory Tests Reference Ranges 2024 [Available from: https://www.abim.org.

22. Akbas BO, Osmanagaoglu MA, Bozkaya H, Ovali E, Ucar F. Prognostic effect of factor V Leiden mutation in pregnant women with preeclampsia, eclampsia and chronic hypertension. 2024.

23. Alsheikh AM, Elsadek AM, Gebreel SA. Prevalence and Clinical Significance of Factor V Leiden Mutation in Egyptian Preeclamptic Women. International Journal of Medical Arts. 2021;3(1):1033–8.

24. Ababio G, Adu-Bonsaffoh K, Abindau E, Narh G, Tetteh D, Botchway F, et al. Effects of factor v Leiden polymorphism on the pathogenesis and outcomes of preeclampsia. BMC Medical Genetics. 2019;20:1–6.

25. Al-Otaiby M, Althnayan R, Binmethem A, AlEnezy RB, Alhadlg MA, Alaqeel A, et al. The prevalence of Factor V Leiden (Arg506Gln) mutation in King Khalid University Hospital patients, 2017–2019. Nagoya Journal of Medical Science. 2021;83(3):407.

26. Jadaon MM. Epidemiology of Activated Protein the Mediterranean Region.

27. Vrtel P, Slavik L, Vodicka R, Stellmachova J, Prochazka M, Prochazkova J, et al. Detection of unknown and rare pathogenic variants in antithrombin, protein C and protein S deficiency using high-throughput targeted sequencing. Diagnostics. 2022;12(5):1060.

28. Kashif S, Kashif MA, Saeed A. The association of factor V leiden mutation with recurrent pregnancy loss. J Pak Med Assoc. 2015;65(11):1169–72.

29. Atiquzzaman M, Alam ASE, Hoque MM, Islam MM, Hamid T, Halim MR, et al. Factor V Leiden Thrombophilia Causing Recurrent Thrombosis. Bangladesh Critical Care Journal. 2021;9(2):104–8.

30. Deveer R, Engin-Ustun Y, Akbaba E, Halisdemir B, Cakar E, Danisman N, et al. Association between pre-eclampsia and inherited thrombophilias. Fetal and pediatric pathology. 2013;32(3):213–7.

31. Madkhaly F, Alshaikh A, Alkhail HA, Alnounou R, Owaidah T. Prevalence of positive factor V Leiden and prothrombin mutations in samples tested for thrombophilia in Saudi Arabia. American Journal of Blood Research. 2021;11(3):255.

32. Vicoveanu P, Dimitriu DC, Gorduza E, Ivona M, Filip C, Socolov D. The association between factor V Leiden, MTHFR C667t/A1298c polymorphisms and pregnancy outcomes. The Medical-Surgical Journal. 2021;125(4):563–9.

33. Akhtar T. Association of Factor V Leiden and Prothrombin Gene Mutation in Male and Placental Mediated Pregnancy Complications in Female with their Risk Factors: CAPITAL UNIVERSITY; 2021.

34. Katz D, Beilin Y. Disorders of coagulation in pregnancy. BJA: British Journal of Anaesthesia. 2015;115(Suppl_2):ii75–ii88.

35. Saleh DK, Al Mudallal SS. Evaluation of Protein C and Protein S in Pregnant Females with Preeclampsia. Iraqi Postgraduate Medical Journal. 2021;20(1).

36. Okoye HC, Eweputanna LI, Okpani AO, Ejele OA. Associations between pre-eclampsia and protein C and protein S levels among pregnant Nigerian women. International Journal of Gynecology & Obstetrics. 2017;137(1):26–30.

37. Mitriuc D, Popuşoi O, Catrinici R, Friptu V. The obstetric complications in women with hereditary thrombophilia. Medicine and pharmacy reports. 2019;92(2):106.

38. Kristoffersen AH, Petersen PH, Røraas T, Sandberg S. Estimates of within-subject biological variation of protein C, antithrombin, protein S free, protein S activity, and activated protein C resistance in pregnant women. Clinical chemistry. 2017;63(4):898–907.

39. Mintaah S, Anto EO, Boadu WIO, Sackey B, Boateng LA, Ansah E, et al. Coagulation Factors and Natural Anticoagulants as Surrogate Markers of Preeclampsia and Its Subtypes: A Case–Control Study in a Ghanaian Population. Clinical and Applied Thrombosis/Hemostasis. 2023;29:10760296231204604.

40. Jung YW, Park DB, An SJ, Chung SM, Kang BH, Yoo HJ, et al. Changes in Protein C and Protein S Activities and the Association with Adverse Pregnancy Outcomes in Pregnant Korean Women. Laboratory Medicine Online. 2024;14(2):82–9.

41. Ahmed EM, Hassanen RH, Abbas AM, Kalaf SA. Predict Risk Factors of Preeclampsia Among Pregnant Women Attended Antenatal Clinic at Assiut University Hospital. Assiut Scientific Nursing Journal. 2018;6(14):145–56.

42. Khidri FF. Various presentations of preeclampsia at tertiary care hospital of Sindh: A Cross-Sectional Study. Current Hypertension Reviews. 2020;16(3):216–22.

43. Asres A, Tilahun A, Waji S, Adella G, Addissie A. Past hormonal contraceptive use and pre-eclampsia among pregnant women in Northwest Ethiopia: a case-control study. Authorea Preprints. 2020.

44. Gunaratne MD, Thorsteinsdottir B, Garovic VD. Combined oral contraceptive pill-induced hypertension and hypertensive disorders of pregnancy: shared mechanisms and clinical similarities. Current hypertension reports. 2021;23:1–13.

